# Association of Trends in Child Undernutrition and Implementation of the National Rural Health Mission in India: A Nationally Representative Serial Cross-Sectional Study on Data from 1992 – 2015

**DOI:** 10.1101/2022.02.28.22271596

**Authors:** Apurv Soni, Nisha Fahey, Zulfiqar A. Bhutta, Wenjun Li, Tiffany Moore Simas, Somashekhar Nimbalkar, Jeroan Allison

## Abstract

**Background:** India launched the National Rural Health Mission (NRHM) in 2005 to strengthen its primary healthcare system in high-focus and northeast focus states. One of the NRHM objectives was to reduce child undernutrition in India.

**Methods and Findings:** We used data from 1992, 1998, 2005, and 2015 National Family Health Surveys of India to evaluate trends in child undernutrition prevalence before and after NRHM and across different categories of focus states. Stunting, Wasting, and Comprehensive Index of Anthropometric Failure (CIAF) was assessed using WHO growth curves to assess chronic, acute, and overall undernutrition. The study included 187,452 children aged 3 years or under. Survey-weighted and confounder-adjusted average annualized reduction rates (AARR) and predicted probability ratios were used to assess trends and socioeconomic disparities for child undernutrition, respectively. Nationwide, the prevalence of all types of undernutrition decreased from 1992 to 2015. However, the trends varied before and after NRHM implementation and differentially by focus states. After NRHM, acute undernutrition declined more rapidly among high-focus states (AARR 1.0%) but increased in normal-focus states (AARR -1.9% per year; p<0.001). In contrast, the prevalence of chronic undernutrition declined more rapidly (AARR 1.6%) in the normal focus states in comparison to high-focus states (0.3%; p=0.01). Income and caste-based disparities in acute undernutrition decreased but did not disappear after the implementation of the NRHM. However, similar disparities in prevalence of chronic undernutrition appear to be exacerbated after the implementation of the NRHM. Major limitations of this study include the observational and cross-sectional design, which preclude our ability to draw causal inferences.

**Conclusion:** Our results suggests that NRHM implementation might be associated with improvement in wasting (acute) rather than stunting (chronic) forms of undernutrition. Strategies to combat undernutrition equitably, especially in high-focus states are needed.

**AUTHOR SUMMARY:** *Why was this study done?:*

- India has the largest number of undernourished children of any country worldwide
- India’s government launched a public health program called National Rural Health Mission (NRHM) that had a focus on improving maternal and child health
- Impact of the NRHM has been studied on infant mortality rate but nationwide estimates of association of NRHM with child undernutrition rates remains unelucidated

*What did the researchers do and find?:*

- We carried out a cross-sectional study based on nationally representative serial data gathered through the National Family Health Surveys in 1992, 1998, 2005, and 2015 on a total 187,452 children, which represented national population of children aged 3 years and under residing in India in the respective survey years
- The study reports that the prevalence of undernutrition and trends in its changes before and after implementation of NRHM differed across different indicators of undernutrition.
- The prevalence of wasting (acute undernutrition) declined at a faster rate in the post-NRHM period among the states where NRHM was prioritized while the prevalence of acute undernutrition increased during the post-NRHM where NRHM was not implemented. Conversely, prevalence of stunting (chronic undernutrition) declined at a faster rate among the normal focus states in the post-NRHM period in comparison to high and northeast focus states.
- Socioeconomic disparities for chronic and overall undernutrition persisted or widened over time. We observed a decline in wealth, maternal-education, rural, and caste-based disparities for acute undernutrition after the implementation of the NRHM.

*What do these findings mean?:*

- Policies enacted under the NRHM may be associated with a reduction in acute form of child undernutrition
- Policies enacted under the NRHM may be associated with modest reduction in wealth, education, or caste-based disparities for acute form of child undernutrition, however, the disparities for chronic undernutrition among children were not associated by the NRHM

## BACKGROUND

In 2016, one in four children less than 5 years old (∼155 million) worldwide experienced stunted growth, an indicator of chronic and intergenerational undernutrition, and approximately one in 10 (∼52 million) suffered from wasting, an indicator of acute undernutrition [1]. Child undernutrition remains a serious public health problem, especially in low- and middle-income countries, where two-thirds of stunted children and three-fourths of wasted children reside [1]. Reduction of child undernutrition has been at the forefront of the world’s development goals for more than a decade, with Goal 1 of the United Nations’ 2005 Millennium Development Goals aiming to reduce the number of underweight children by one half of the prevalence levels in 1990 by 2015 [2]. India, one of the signatories of Millennium Development Goals, failed to meet this goal.

A disproportionate share of the world’s undernourished child population reside in India [1,3]. Despite its economic growth, the rate of institutional deliveries and coverage of maternal and newborn care practices remains among the lowest in South Asia [4,5]. Additionally, there were tremendous interstate and rural-urban disparities in maternal and child health indicators throughout India [6,7]. In response, the Indian government launched a flagship national health program called National Rural Health Mission (NRHM), which had a special focus on maternal and child health in the rural regions of 18 states: 10 large and impoverished states were considered as high focus while 8 states in northeastern region were considered as the northeast focus because of poor existing healthcare infrastructure in those states [8,9]. Under the NRHM, the Indian government invested more than $12.1 billion from 2005 to 2012 to strengthen public health infrastructure and workforce in rural India [9,10]. Through the NRHM, India developed a workforce of approximately 1 million frontline health workers called Accredited Social Health Activist (ASHA: *hope*), who are responsible for tracking pregnant women, young mothers, and children under the age of five. These workers also facilitate healthcare delivery by accompanying the beneficiaries to scheduled clinical and immunization visits. While one study found that the NRHM has led to a reduction in urban-rural disparities in the infant mortality rate,[11] there is a lack of evidence about the impact of NRHM programs and policies on the magnitude of child undernutrition in India.

This study investigates changes in the prevalence of child undernutrition in India from 1992 to 2015-16 and assesses differential change in the rate of child undernutrition reduction, pre and post-NRHM, for states based on their focus classification. The secondary study objective was to examine whether socio-economic inequalities persisted or declined after the implementation of the NRHM and whether this reduction was greater among high-focus states in comparison with normal-focus states.

## METHODS

### Study Setting and Data

This study analyzes data from 1992, 1998, 2005, and 2015 cross-sectional surveys of the population of India called National Family Health Survey (NFHS) [4,12–14]. These surveys are conducted using standard model questionnaires designed for Demographic and Health Surveys (DHS), which were developed by the United States Agency for International Development (USAID) and widely used in low- and middle-income countries. In India, these surveys use census data to perform multi-stage cluster sampling that results in a representative sample of the Indian population. Details of these surveys have been described elsewhere [4,12–14]. Data are available to registered users through the DHS website (www.dhsprogram.com) and pooled data are available through Integrated Public Use Microdata Series-DHS, a grant-based initiative to promote microdata analyses of DHS datasets[15]. This study did not have a pre-specified analysis plan prior to collection of the data. Separate questionnaires are administered to the head of household, women aged 15-49 years, and men aged 15-54 years who stayed in the household the night before the household interview was performed. Women of reproductive age are also asked to provide information about their children. These surveys collect data on a wide range of topics, including women’s reproductive history, household characteristics, anthropometric measurements of children and women of reproductive age, and maternal and child health indicators. Because child anthropometry was only measured for children under three years old in 1998, and only among those who were born to women in the household, we limited our sample from other years to children who meet these criteria. Additionally, we excluded children who were deceased at the time of the survey and those with missing anthropometric or age data. The original sample size, number of children meeting the exclusion criteria, and the final analytic samples are shown in the *Figure 1*.

**Figure 1:**
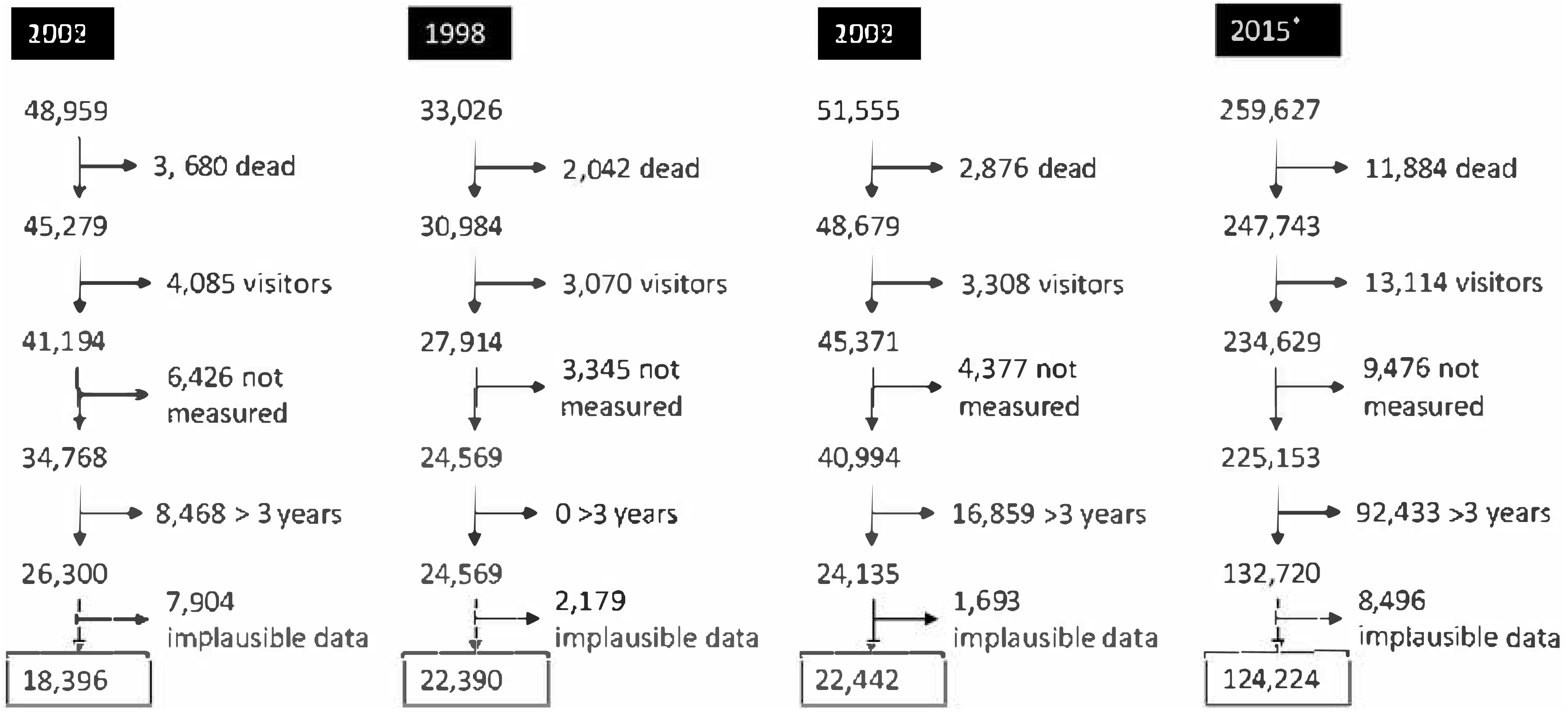
Flowchart demonstrating exclusions and final analytic sample from the National Family Health Surveys in India included in this study. * 2015 survey sample size was increased to derive estimates at the district levels Characteristics of the cohort did not markedly change intra- and inter-surveys based on the different exclusion criteria (results not shown)

### Variables

#### Outcome

The outcome was measured using three indicators to characterize chronic, acute, and overall burden of child undernutrition separately. We used the World Health Organization (WHO) standards to compute the Z scores for weight-for-age, weight-for-height, and height-for-age [16]. A child was considered to be chronically undernourished or stunted if his/her height-for-age Z score was below -2 and acutely undernourished or wasted if his/her weight-for-height Z score was below -2. Using recommendations of a WHO working paper, we operationalized overall burden of undernutrition as Composite Index of Anthropometric Failure – CIAF if a child was either stunted, wasted, or underweight (weight-for-age Z score below -2) [17–19]. Although we calculated underweight status of the children in the process of classifying CIAF status, we do not report statistics based on underweight because it does not distinguish between acute and chronic form of undernutrition and does not completely represent the overall burden of child undernutrition [20].

#### Exposure

We defined the time from 1992 to 2005 as the pre-NRHM period and 2005 to 2012 as the post-NRHM period. Because the 2005 NFHS was primarily conducted before July 2005 and the NRHM program was not implemented until the end of 2005, we considered survey data from 2005 to be in the pre-NRHM period. State focus classification was based on the official list provided by the Indian government [9]. To assess socioeconomic inequalities, we considered wealth quintile, level of maternal education, caste, rural/urban residence, and sex of the child as secondary exposure variables. Household wealth was operationalized using quintiles of a household assets score created using principal components analysis on items related to possession of assets [21]. This method has been found to correlate highly with income and expenditure data in India [22]. Maternal education was operationalized into three categories: no formal education (0 years), primary (1-5 years), secondary and higher (6+ years) levels of education. We categorized the caste variable as General, Other Backwards Class, and Scheduled Caste or Tribe. Rural or urban residence was based on the census definition of India and Sex of the child was based on maternal report.

#### Potential Confounding Variables

Previous analyses of child undernutrition using DHS data from other countries assessed trends and inter-country comparisons after accounting for child’s age, sex, rural residence, birth order, religion, and maternal age [23,24]. We also considered month of survey as a confounder because evidence has shown that weight is influenced by seasonality.

### Data Analysis

The weighted prevalence of child undernutrition indicators over time and according to subgroups of exposure categories was estimated by accounting for clustered sampling design and using survey weights. Because of the asymmetrical timeline of surveys, we used the UNICEF formula for estimating annualized changes as average annual reduction rates (AARRs): 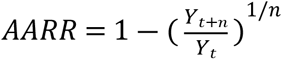 where Y is the adjusted prevalence of the childhood nutrition indicator at time (t) and (n) is the number of years between (t) and (t+n) [24,25]. Adjusted prevalence of different undernutrition indicators was estimated by performing the inverse logit transformation of coefficients from separate survey-weighted multivariable logistic regression models which included an interaction term for time and focus state classification. The multivariable model also included month of survey as a dummy variable, caste, and religion. Other confounders were not included because they might lie in the pathway of NRHM programs and policies’ impact on child nutrition. For example, some of the NRHM interventions focused on optimal birth spacing, child-bearing age, and advocacy of female child. Thus, including child sex, birth order, rural location and maternal age in the model would lead to over-adjustment. Similarly, household and structural determinants were addressed by the NRHM and affiliated government programs that focused on improving the health and well-being of reproductive aged women, thus they were excluded as confounders from the primary analysis. Point estimates for AARR, differences in AARR, and differences of differences in AARR were calculated with accompanying 95% Confidence Interval (CI) using the Delta method that employs Taylor series linearization, which is the same methodology used by the WHO-UNICEF Technical Expert Advisory Group on Nutrition Monitoring.[26] Socio-economic disparities were represented using the predicted probability ratio of adjusted estimates after accounting for all potential confounders. In order to emphasize inequalities, predicted probability ratios were calculated for extreme categories for household wealth, maternal education, and caste because they had more than two categories. Due to small sample size, socio-economic disparities for northeast focus states were not reliable and not estimated. All analyses were performed in Stata 15 using svy prefix to account for the complex survey data structure.

### Ethics

The University of Massachusetts Medical School IRB reviewed the protocol for the secondary data analyses presented in this manuscript. This study was deemed exempt from full review because the data contained no personal identifiable information on survey participants.

## RESULTS

Among the total of 393,167 eligible children included in the NFHS surveys from 1992 to 2015, 187,452 met the criteria for inclusion in the present study (*Fig 1*). *Table 1* presents the weighted distribution of variables of interest in this study. Most of the children surveyed resided in high focus states and less than 5% lived in northeast focus states. The proportion of children living in the poorest wealth quintile increased over time while the proportion living in the richest wealth quintile decreased. There was a dramatic increase in maternal education over the years. In 1992, nearly two-thirds reported attending zero years of school while in 2015, approximately one quarter of mothers reported no formal education. The proportion of children living in rural regions declined over time as did the proportion of female children. All observed trends were consistent across different categories of focus states (*S1 Table1*). Children living in normal focus states had higher socioeconomic status in comparison to children living in high or northeast focus states. These trends were also consistent within the excluded sample as demonstrated in supplementary tables (*S1 Table 2*).

**Table 1:**
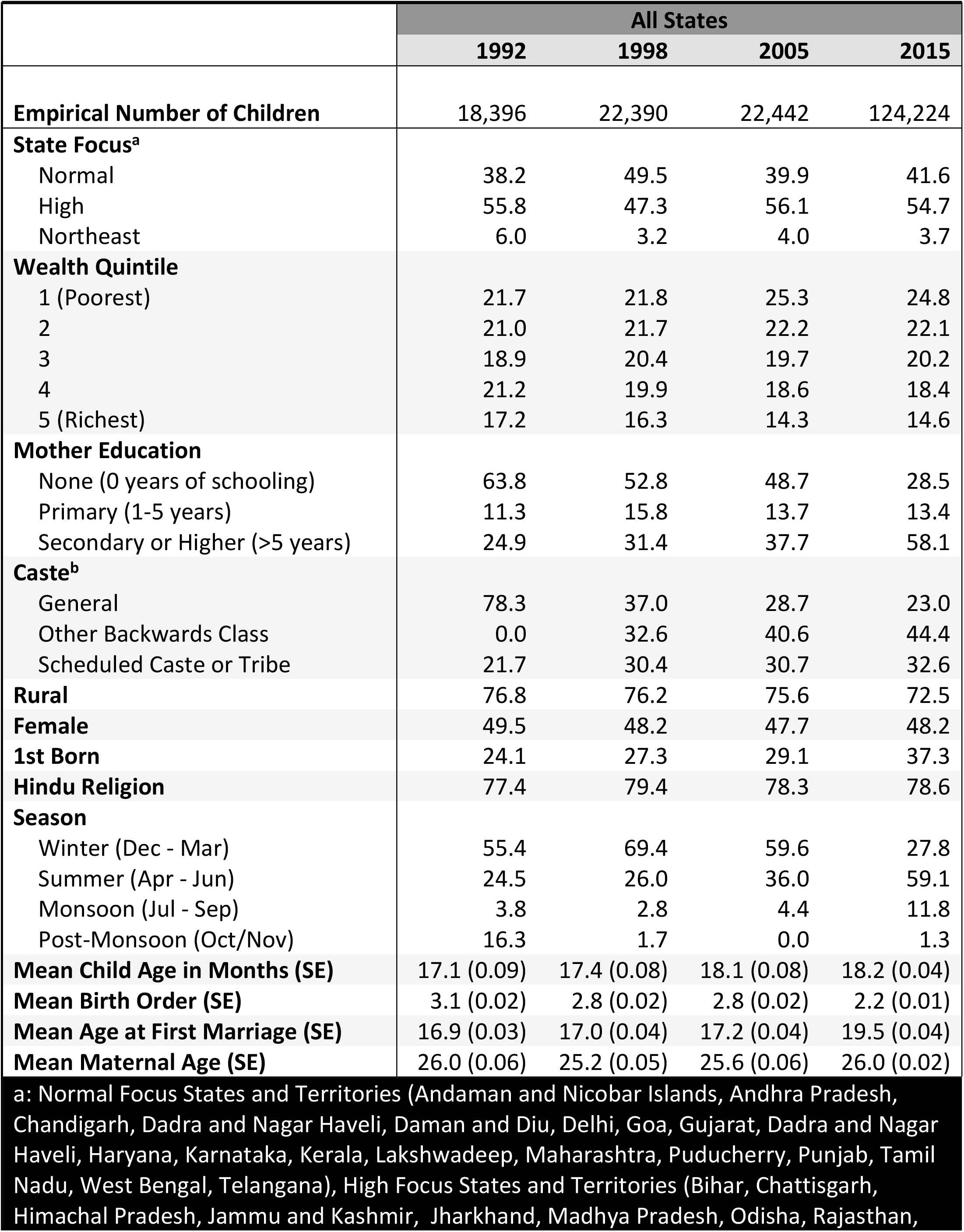
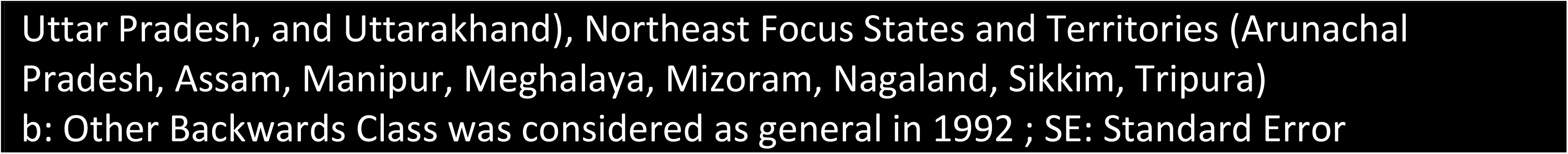
Weighted distribution of eligible children under the age of 3 measured for undernutrition in Indian National Family Health Surveys from 1992 to 2015.

|  |  |  |  |  |
| --- | --- | --- | --- | --- |
|  | **All States** |  |  |  |
|  | **1992** | **1998** | **2005** | **2015** |
| **Empirical Number of Children** | 18,396 | 22,390 | 22,442 | 124,224 |
| **State Focus<sup>a</sup>** |  |  |  |  |
| Normal | 38.2 | 49.5 | 39.9 | 41.6 |
| High | 55.8 | 47.3 | 56.1 | 54.7 |
| Northeast | 6.0 | 3.2 | 4.0 | 3.7 |
| **Wealth Quintile** |  |  |  |  |
| 1 (Poorest) | 21.7 | 21.8 | 25.3 | 24.8 |
| 2 | 21.0 | 21.7 | 22.2 | 22.1 |
| 3 | 18.9 | 20.4 | 19.7 | 20.2 |
| 4 | 21.2 | 19.9 | 18.6 | 18.4 |
| 5 (Richest) | 17.2 | 16.3 | 14.3 | 14.6 |
| **Mother Education** |  |  |  |  |
| None (0 years of schooling) | 63.8 | 52.8 | 48.7 | 28.5 |
| Primary (1-5 years) | 11.3 | 15.8 | 13.7 | 13.4 |
| Secondary or Higher (>5 years) | 24.9 | 31.4 | 37.7 | 58.1 |
| **Caste<sup>b</sup>** |  |  |  |  |
| General | 78.3 | 37.0 | 28.7 | 23.0 |
| Other Backwards Class | 0.0 | 32.6 | 40.6 | 44.4 |
| Scheduled Caste or Tribe | 21.7 | 30.4 | 30.7 | 32.6 |
| **Rural** | 76.8 | 76.2 | 75.6 | 72.5 |
| **Female** | 49.5 | 48.2 | 47.7 | 48.2 |
| **1st Born** | 24.1 | 27.3 | 29.1 | 37.3 |
| **Hindu Religion** | 77.4 | 79.4 | 78.3 | 78.6 |
| **Season** |  |  |  |  |
| Winter (Dec - Mar) | 55.4 | 69.4 | 59.6 | 27.8 |
| Summer (Apr - Jun) | 24.5 | 26.0 | 36.0 | 59.1 |
| Monsoon (Jul - Sep) | 3.8 | 2.8 | 4.4 | 11.8 |
| Post-Monsoon (Oct/Nov) | 16.3 | 1.7 | 0.0 | 1.3 |
| **Mean Child Age in Months (SE)** | 17.1 (0.09) | 17.4 (0.08) | 18.1 (0.08) | 18.2 (0.04) |
| **Mean Birth Order (SE)** | 3.1 (0.02) | 2.8 (0.02) | 2.8 (0.02) | 2.2 (0.01) |
| **Mean Age at First Marriage (SE)** | 16.9 (0.03) | 17.0 (0.04) | 17.2 (0.04) | 19.5 (0.04) |
| **Mean Maternal Age (SE)** | 26.0 (0.06) | 25.2 (0.05) | 25.6 (0.06) | 26.0 (0.02) |
| a: Normal Focus States and Territories (Andaman and Nicobar Islands, Andhra Pradesh, Chandigarh, Dadra and Nagar Haveli, Daman and Diu, Delhi, Goa, Gujarat, Dadra and Nagar Haveli, Haryana, Karnataka, Kerala, Lakshwadeep, Maharashtra, Puducherry, Punjab, Tamil Nadu, West Bengal, Telangana), High Focus States and Territories (Bihar, Chattisgarh, Himachal Pradesh, Jammu and Kashmir, Jharkhand, Madhya Pradesh, Odisha, Rajasthan, |  |  |  |  |

Uttar Pradesh, and Uttarakhand), Northeast Focus States and Territories (Arunachal Pradesh, Assam, Manipur, Meghalaya, Mizoram, Nagaland, Sikkim, Tripura) b: Other Backwards Class was considered as general in 1992 ; SE: Standard Error

Across India, the prevalence of acute, chronic, and overall undernutrition decreased during the years under study (*Fig 2A*). Chronic undernutrition declined at a 0.8% faster annual rate after the implementation of the NRHM program (95% CI: 0.4 – 1.2). In contrast, prevalence of acute undernutrition increased during the post-NRHM period. We did not observe a discernible change in the AARR between post-NRHM and pre-NRHM period for overall undernutrition. There was heterogeneity in these observations among the subgroups of states. Among normal focus states, chronic undernutrition declined at a 1.6% faster rate during the post-NRHM in comparison to pre-NRHM period, whilst prevalence of acute undernutrition increased (*Fig 2B*). Among high and northeast focus states, prevalence of acute undernutrition declined at a faster rate during the post-NRHM period in comparison to pre-NRHM period while no statistically significant difference was found in the change of rate of decline for chronic or overall undernutrition (*Fig 2C and 2D*). These changes in AARR after the implementation of the NRHM were significantly different between normal focus states and the states where NRHM was implemented i.e. high and northeast focus (*Table 2*). Although not the focus of this study, changes in trends of underweight and double-burden of acute (wasting) and chronic (stunting) undernutrition is described Supplementary Table 3 (S1 Table 3) Socioeconomic disparities of child undernutrition from 1992 to 2015 are presented using the predicted probability ratio between two extreme categories in *Fig 3*. A predicted probability ratio of 1 indicates that there are no disparities based on the variable of interest, while a predicted probability ratio >1 is indicative of a disparity such that children in the lower socioeconomic category have an increased prevalence in comparison with children in the higher socioeconomic category. Socioeconomic disparities of chronic undernutrition were exacerbated between 1992 and 2015 and no decline in disparities was observed during the post-NRHM period. In contrast, disparities based on wealth, maternal education, caste, and rural residence for acute undernutrition declined but did not disappear during the post-NRHM period. Socioeconomic disparities for CIAF have persisted, despite some attenuation in wealth-based disparity during the post-NRHM period. Trends in disparity did not differ dramatically between high focus and normal focus states except in a few notable cases (*Fig 4*). Maternal education-based disparities in chronic undernutrition declined during the post-NRHM period among normal focus states. In contrast, declines in disparities for chronic undernutrition were not observed among high focus states. Wealth and maternal education-based disparities in acute undernutrition declined during the post-NRHM period for normal and high focus states; the magnitude of decline was greater among the high focus states.

**Figure 2:**
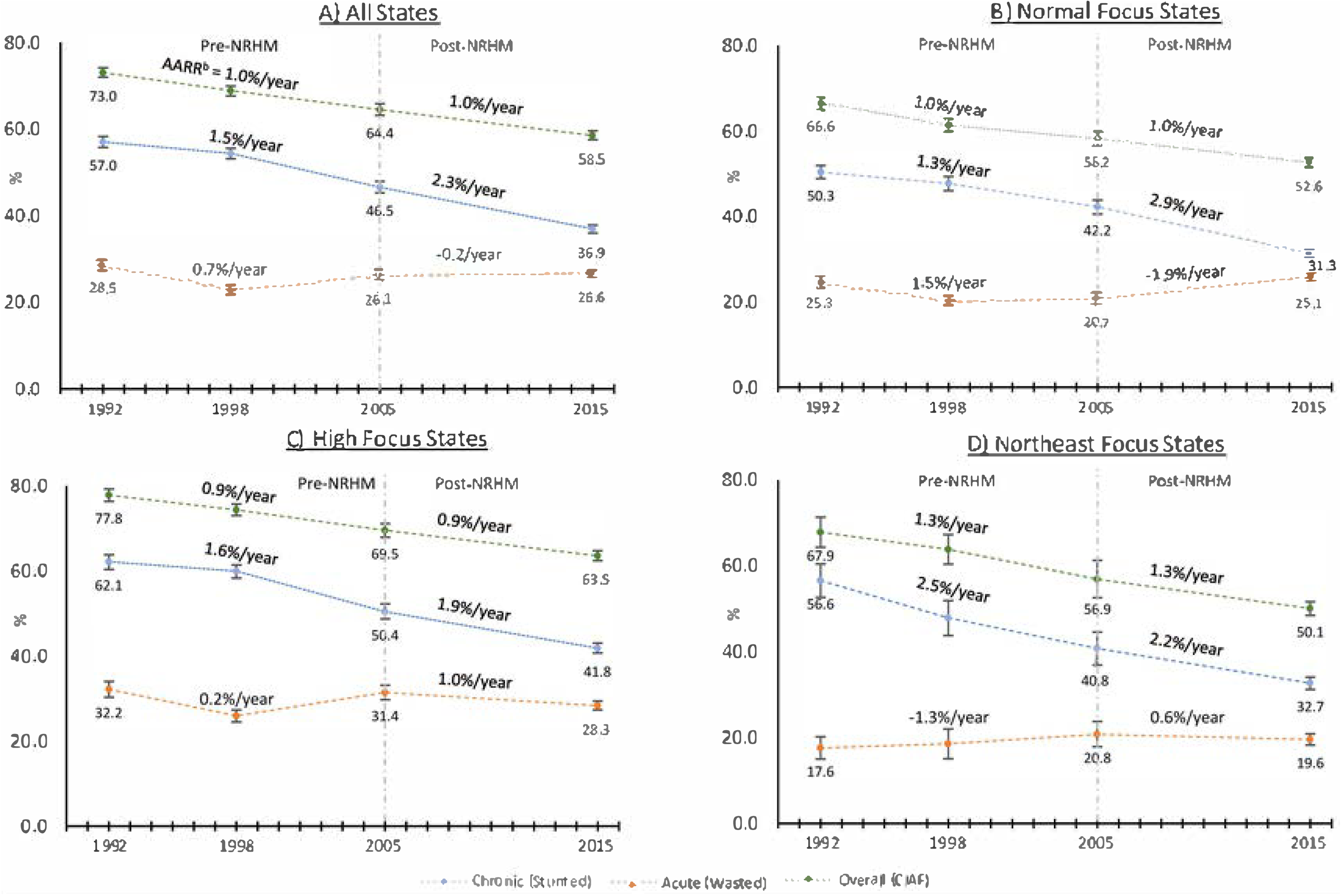
Change in the prevalence^a^ of undernutrition among Indian children aged 3 or less. Data from the 1992, 1998, 2005, and 2015 National Family Health Surveys. a: prevalence adjusted for caste, religion, and month of survey b: Average Annualized Rate of Reduction NRHM: National Rural Health Mission, a national program implemented in 2005 to improve maternal and child health among High focus and Northeast focus states CIAF: Composite Index of Anthropometric Failure considers a child to be undernourished if the child is either stunted, wasted, or underweight

**Table 2:** Differences of the differences between focus states groups for Average Annualized Reduction Rate changes in the post-NRHM (2005-15) versus pre-NRHM periods (1992-2005) based on results from the Indian National Family Health Surveys.

|  | Chronic |  |  |  | Acute |  |  |  | Overall |  |  |  |
| --- | --- | --- | --- | --- | --- | --- | --- | --- | --- | --- | --- | --- |
| | $\Delta$ | LCI | UCI | p | $\Delta$ | LCI | UCI | p | $\Delta$ | LCI | UCI | p |
| (High Focus) - (Normal Focus) | -1.3 | -2.2 | -0.4 | <0.001 | 4.3 | 2.8 | 5.7 | <0.001 | 0.0 | -0.5 | 0.7 | 0.81 |
| (Northeast Focus) - (Normal Focus) | -1.9 | -3.7 | -0.1 | 0.04 | 5.4 | 2.4 | 8.3 | <0.001 | 0.0 | -1.5 | 1.4 | 0.96 |
| (High Focus) - (Northeast Focus) | 0.6 | -1.2 | 2.3 | 0.53 | -1.1 | -4.0 | 1.7 | 0.45 | 0.1 | -1.3 | -1.5 | 0.88 |

Chronic undernutrition was defined by stunting, acute by wasting, and overall by Composite Index of Anthropometric Failure (stunted, wasted, or underweight)
All estimates were adjusted for month of survey as well as religion and caste of the child's household

**Figure 3:**
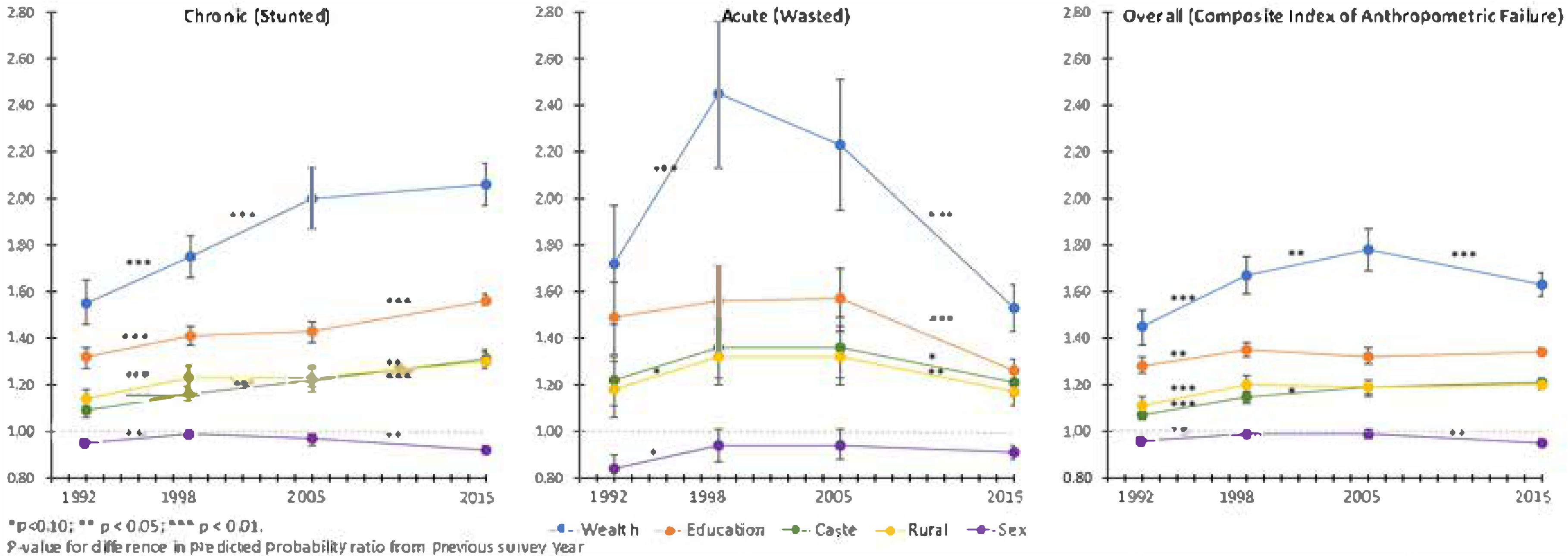
Trends in disparities for undernutrition among Indian children aged 3 or less based on wealth, maternal education, caste, location of residence, and child sex. Data from the 1992, 1998, 2005, and 2015 National Family Health Surveys. Predicted probability ratio for wealth (lowest quintile/highest quintile), maternal education (none/secondary or higher), caste (scheduled caste or tribe/general), location of residence (rural/urban), and child sex (female/male) were estimated after mutually adjusting for each other as well as child’s age, birth order, religion, and maternal age.

**Figure 4:**
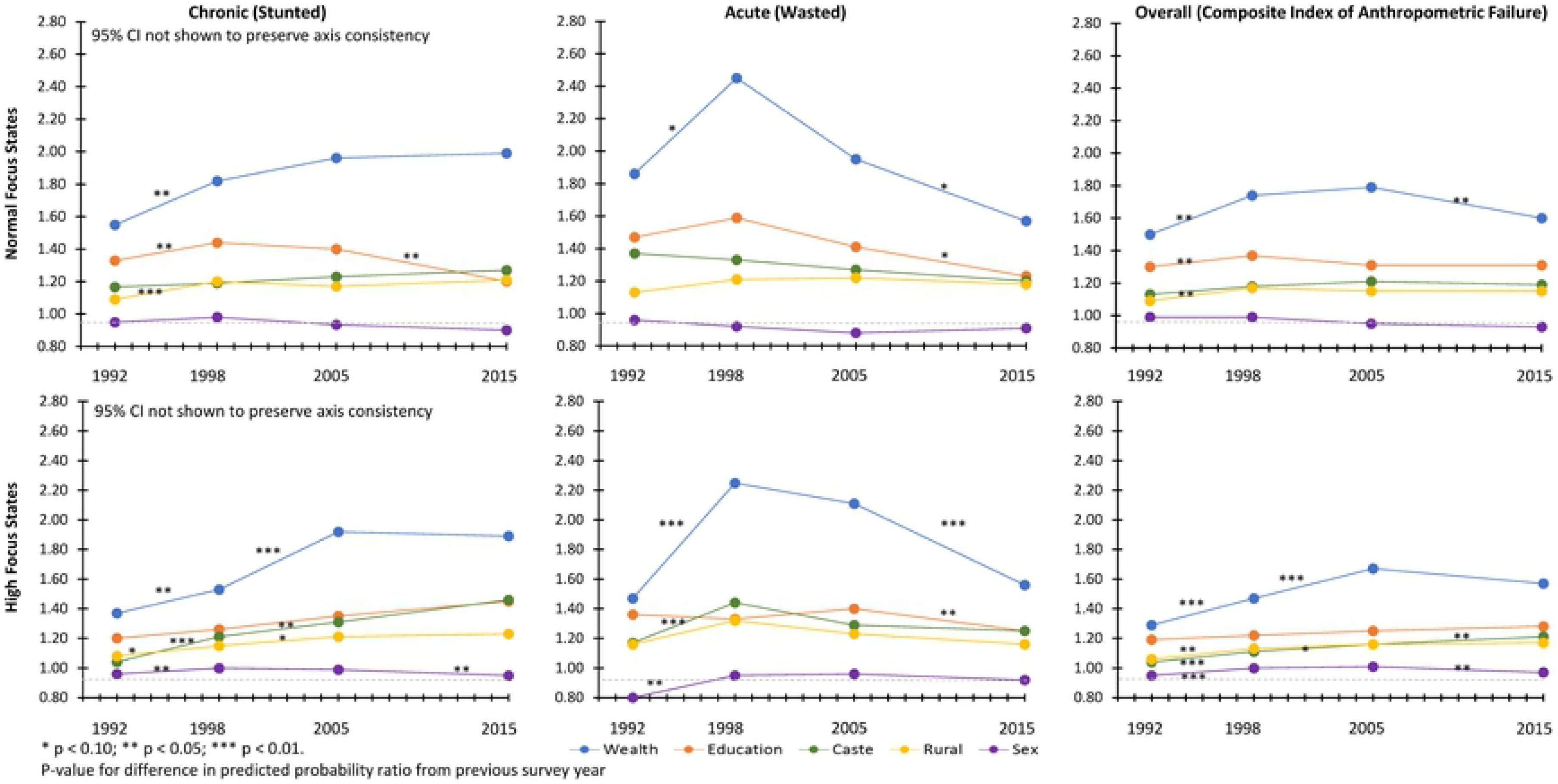
Trends in socio-economic disparities for child undernutrition among Normal Focus and High Focus states. Data from the 1992,1998, 2005, and 2015 National Family Health Surveys. Predicted probability ratio for wealth (lowest quintile/highest quintile), maternal education (none/secondary or higher), caste (scheduled caste or tribe/general), location of residence (rural/urban), and child sex (female/male) were estimated after mutually adjusting for each other as well as child’s age, birth order, religion, and maternal age. Northeast focus states not shown because prevalence ratio were not estimable due to small sample size

## DISCUSSION

Our study used nationally representative cross-sectional data in India during the 4 survey years of 1992, 1998, 2005, and 2015 on eligible children under the age of three to examine trends in child undernutrition within the context of the National Rural Health Mission that was implemented in 2005 among the high focus and northeast focus states in India. Our analyses reveal three key findings. First, the prevalence of undernutrition and trends in its changes differed across different indicators of undernutrition. Second, the prevalence of acute undernutrition declined at a faster rate in the post-NRHM period among the states where NRHM was prioritized while the prevalence of acute undernutrition increased during the post-NRHM where NRHM was not implemented. Conversely, prevalence of chronic undernutrition declined at a faster rate among the normal focus states in the post-NRHM period in comparison to high and northeast focus states. Third, socioeconomic disparities for chronic and overall undernutrition persisted or widened over time. We observed a decline in wealth, maternal-education, rural, and caste-based disparities for acute undernutrition after the implementation of the NRHM. These results have important implications for the Indian government as it continues to expand its efforts to combat child undernutrition.

Conventionally, prevalence of child undernutrition is assessed using height-for-age (stunting), which is used by the WHO to set targets for its Millennium and Sustainable Development Goals [27,28]. Our findings suggest that focusing on any one indicator underestimates the overall prevalence of undernutrition, which is better captured using CIAF [19]. Our results also suggest a sustained reduction in children who meet the criteria for stunted growth since 1992, especially in the post-NRHM period when prevalence decreased by more than 2% per year. However, the overall burden of child undernutrition, measured by CIAF, has decreased by 1% from 1992. Moreover, the rate of reduction in CIAF in the post-NRHM period was not higher than in the pre-NRHM period. This finding that stunting overestimates the changes in undernutrition is consistent with analyses of changes in the prevalence of child undernutrition indicators from seven low- and middle-income countries, which found contradictory findings [20]. As an example, the relative change in the prevalence of stunting and wasting among Zimbabwean children under the age of 5 increased by 24.5% and 32.7%, respectively, from 1994 to 1999, while the prevalence of underweight children decreased by 10.2% [20]. Conventional indices of stunting and wasting correspond to distinct biological phenomena and must be considered in informing interventions, but policy impact evaluation should also consider CIAF to more comprehensively assess improvement in the overall nutritional status of children and more effectively allocate necessary resources.

The NRHM was implemented in 2005 among the rural regions of the 18 high-focus states to reduce geographical disparities in maternal and child health [8,9,29,30]. We found that the prevalence of acute undernutrition decreased at a more rapid rate during the post-NRHM period among children living in high focus states in comparison to normal focus states, supporting our original hypothesis. These findings may be explained by the services provided by the NRHM, which focused on improving antenatal care, rates of institutional delivery, and healthcare access for mother and the child [31]. These services are more likely to prevent acute forms of undernutrition by improving health and preventing child morbidity. Additionally, nutritional rehabilitation centers instituted under the NRHM can help children recover from acute malnutrition. Recent analysis found that NRHM was associated with a reduction in wealth-based inequities in maternal health services [32]. Similarly, our finding that socioeconomic disparities for acute undernutrition declined suggests that NRHM has been effective in combating acute malnutrition among the disadvantaged populations.

Contrary to our hypothesis, we found that the prevalence of stunting decreased with a higher rate among normal focus states in comparison to high focus states during the post-NRHM period. One plausible reason for the unexpected trends is that determinants of chronic nutrition are less influenced by the NRHM policies, at least on a short-term basis. An analysis of determinants of child stunting using nationally representative data from Afghanistan, Bangladesh, India, Nepal, and Pakistan showed that the three strongest correlates of child stunting were maternal height, household wealth, maternal BMI, minimum dietary diversity, maternal education, and age at marriage [33]. It is possible that improved access to healthcare services, may help increase maternal height and BMI as well as reduce economic burden on the families, thereby increasing household wealth. However, meaningful changes in these factors may not manifest in the observed post-NRHM period. Therefore, the higher rate of reduction in the normal focus states may be a reflection of the overall status of women in these states where 74% of mothers had received secondary or higher level of education in comparison to only 45.4% in high focus states. An important implication of these results is that India needs to invest in programs that more directly improve the status of women in high focus states to reduce the prevalence of children with stunted growth.

Since 2016, India has launched two nationwide programs to improve maternal and child health, with a special focus on child nutrition. Mothers Absolute Affection (MAA: *mother*) is a program intended to enhance breastfeeding and other young child feeding practices through a multimedia information campaign and by leveraging the ASHA workforce to conduct regular meetings with young mothers from their community [34]. Prime-minister’s Overarching Scheme for Holistic Nourishment (POSHAN: *Nutrition*) or National Nutrition Mission (NNM) is a three-year 1.5 billion U.S. dollar initiative that aims to use mobile technology to integrate siloed efforts to improve immediate and underlying determinants of child undernutrition.[35] The goal of POSHAN is to reduce the prevalence of stunting and underweight by 2% per year and achieve a prevalence of less than 25% for both indicators by 2025. Both of these programs represent a step in the right direction, but their mission guidelines indicate an implementation strategy based on state and district-based priority. As our findings on the impact of the NRHM shows, careful consideration for effective allocation of resources is necessary to reduce disparities and limit unintended effects. In the context of rising wealth inequality in India, prioritizing implementation based on high focus states puts the impoverished and disenfranchised population in normal states at higher risk as observed in our findings. Therefore, there is a need for a mechanism that more effectively targets vulnerable populations across India. The emergence of mobile technology and existence of a robust workforce of frontline health workers might offer a way forward towards this goal.

Our study has several limitations. First, we considered exposure of NRHM policies based on state classification by the Indian government as high focus, northeast focus, or normal focus. Because the implementation of NRHM program is at the discretion of the state government to allow for contextual modification, interstate variations might exist. Thus, our approach overlooks the underlying heterogeneity within states of a particular focus classification. However, this analytical framework has been used by previous evaluation of NRHM program [10,30,32]. Second, NRHM was expanded to National Health Mission (NHM) in 2012 that included urban regions and normal focus states [36]. Therefore, it is possible that our findings reflect an underestimation of the impact of NRHM. However, previous analyses of the NRHM policies showed that measurable differences in health indicators were observed five years after the implementation of NRHM. The magnitude of differences found between high and normal focus states also suggests that an underestimation of the impact of NRHM is unlikely to change the overall findings. Additionally, programmatic guidelines for new and existing programs continue to consider states’ focus classification for their implementation strategy. Third, our study assumes a natural design approach and mimicks an interrupted time series design but there are limited timepoints for nationally representative estimation of child undernutrition. Other datasets such as District Level Household Surveys provide more frequent assessment of nutrition in certain states but these data do not cover the full geography of India and preclude our ability to have more robust understanding of trends over time within the intervention period. Finally, there is a considerable amount of missing or implausible data. However, the distribution of sociodemographic factors did not vary between the missing and non-missing groups and the missingness pattern did not differentially change between the focus group-states. Considering that this source of data has been widely used in literature and informs policy making by the Indian government, we believe that it is currently the best available data for this analysis.

In conclusion, our results suggest differential improvements in chronic and acute undernutrition for normal and high-focus states after the implementation of NRHM. Moreover, these differential improvements were also reflected in improvement of disparities for acute undernutrition but exacerbation of disparities for chronic undernutrition Our findings suggest that NRHM programs and associated investment in perinatal health might be associated with a decline in acute forms of undernutrition. However, there is a need for India to more effectively focus its interventions on vulnerable populations to reduce child undernutrition-based disparities.

## Data Availability

All data used in this study are available publicly and free of cost to registered users through the DHS website (www.dhsprogram.com) and Integrated Public Use Microdata Series-DHS, a grant-based initiative to promote microdata analyses of DHS datasets (www.idhsdata.org/idhs/).

https://www.idhsdata.org/idhs/

## REFERENCES

1. UNICEF. The State of the World’s Children 2016: A fair chance for every child. Geneva; 2016. Available: https://www.unicef.org/publications/files/UNICEF_SOWC_2016.pdf

2. United Nations. The Millenium Development Goals Report 2005. United Nations. 2014.

3. United Nation Children’s Fund. State of the World’s Children 2008. Indian Pediatr. 2008.

4. The Ministry of Health and Family Welfare. National Family Health Survey - 3. Mumbai; 2007. Available: http://rchiips.org/nfhs/nfhs3_national_report.shtml

5. Paul VK, Sachdev HS, Mavalankar D, Ramachandran P, Sankar MJ, Bhandari N, et al. Reproductive health, and child health and nutrition in India: meeting the challenge. Lancet. 2011;377: 332–49. doi:10.1016/S0140-6736(10)61492-4

6. Kumar C, Singh PK, Rai RK. Coverage gap in maternal and child health services in India: assessing trends and regional deprivation during 1992-2006. J Public Health (Oxf). 2013;35: 598–606. doi:10.1093/pubmed/fds108

7. Sanneving L, Trygg N, Saxena D, Mavalankar D, Thomsen S. Inequity in India: the case of maternal and reproductive health. Glob Health Action. 2013;6: 19145. doi:10.3402/gha.v6i0.19145

8. Mudur G. India launches national rural health mission. BMJ. 2005;330: 2005.

9. National Rural Health Mission. National Rural Health Mission (2005-2012)--Mission document. Indian J Public Health. 49: 175–83. Available: http://www.ncbi.nlm.nih.gov/pubmed/16468284

10. Nagarajan S, Paul VK, Yadav N, Gupta S. The National Rural Health Mission in India: Its impact on maternal, neonatal, and infant mortality. Semin Fetal Neonatal Med. 2015;20: 315–320. doi:10.1016/j.siny.2015.06.003

11. Pandey A, Mohan A. The role of national rural health mission in reducing infant mortality rate in India. Int J Heal Gov. 2018; IJHG-09-2018-0044. doi:10.1108/IJHG-09-2018-0044

12. International Institute for Population Sciences. National Family Health Survey(NFHS-4), 2015-16: India. Mumbai, India; 2017. Available: https://dhsprogram.com/pubs/pdf/FR339/FR339.pdf

13. International Institute for Population Sciences, ORC Macro. National Family Health Survey (NFHS-2), 1998-99: India. Mumbai, India; 2000. Available: https://www.dhsprogram.com/pubs/pdf/FRIND2/FRIND2.pdf

14. International Institute for Population Sciences. National Family Health Survey (MCH and Family Planning), India 1992-93. Mumbai, India; 1995. Available: https://dhsprogram.com/pubs/pdf/FRIND1/FRIND1.pdf

15. Boyle EH, King M, Sobek M. IPUMS-Demographic and Health Surveys: Version 4.0 [dataset]. Minneapolis, USA: Minnesota Population Center and ICF International; 2017. p. NICHD Grant Number R01HD069471.

16. de Onis M, Woynarowska B. [WHO child growth standards for children 0-5 years and the possibility of their implementation in Poland]. Med Wieku Rozwoj. 2010;14: 87–94. doi:10.1088/1751-8113/44/8/085201

17. Nandy S, Irving M, Gordon D, Subramanian S V., Smith GD. Poverty, child undernutrition and morbidity: New evidence from India. Bull World Health Organ. 2005;83: 210–6. doi:/S0042-96862005000300014

18. Nandy S, Svedberg P. The Composite Index of Anthropometric Failure (CIAF): An Alternative Indicator for Malnutrition in Young Children. Handbook of Anthropometry. New York, NY: Springer New York; 2012. pp. 127–137. doi:10.1007/978-1-4419-1788-1_6

19. Svedberg P. Poverty and Undernutrition. New Delhi, India: Oxford University Press; 2000. doi:10.1093/0198292686.001.0001

20. Nandy S, Miranda JJ. Overlooking undernutrition? Using a composite index of anthropometric failure to assess how underweight misses and misleads the assessment of undernutrition in young children. Soc Sci Med. 2008;66: 1963–6. doi:10.1016/j.socscimed.2008.01.021

21. Rutstein SO, Staveteig S. Making the Demographic and Health Surveys Wealth Index comparable. DHS Methodol Reports. 2014.

22. Filmer D, Pritchett LH. Estimating Wealth Effects without Expenditure Data-or Tears: An Application to Educational Enrollments in States of India. Demography. 2001. doi:10.2307/3088292

23. Subramanyam MA, Kawachi I, Berkman LF, Subramanian S V. Socioeconomic inequalities in childhood undernutrition in India: analyzing trends between 1992 and 2005. PLoS One. 2010;5: e11392. doi:10.1371/journal.pone.0011392

24. Krishna A, Mejí a-Guevara I, McGovern M, Aguayo V, Subramanian S V. Trends in inequalities in child stunting in South Asia. Matern Child Nutr. 2017. doi:10.1111/mcn.12517

25. UNICEF. How to calculate average annual rate of reduction (AARR) of underweight prevalence. Geneva, Switzerland; 2007. Available: https://data.unicef.org/wp-content/uploads/2015/12/Technical_Note_AARR_185.pdf

26. World Health Organization and the United Nations Children’s Fund (UNICEF) Technical Expert Advisory Group on Nutrition Monitoring (TEAM). Methodology for monitoring progress towards the global nutrition targets for 2025. 2017.

27. WHO. Physical status: the use and interpretation of anthropometry. Report of a WHO Expert Committee. World Health Organ Tech Rep Ser. 1995;854: 1–452. doi:854

28. de Onis M, Garza C, Victora CG, Onyango AW, Frongillo EA, Martines J. The WHO Multicentre Growth Reference Study: planning, study design, and methodology. Food Nutr Bull. 2004;25: S15–26. doi:10.1177/15648265040251S103

29. Dhingra B, Dutta AK. National rural health mission. Indian J Pediatr. 2011;78: 1520–1526. doi:10.1007/s12098-011-0536-4

30. Husain Z. Health of the National Rural Health Mission. Medicine (Baltimore). 2012;xlvi: 53–60.

31. National Rural Health Mission. Operational Guidelines on Facility Based Management of Children with Severe Acute Malnutrition. 2011. Available: http://www.cmamforum.org/Pool/Resources/Operational-guidelines-on-facility-ased-management-of-children-with-severe-acute-malnutrition-India-2011.pdf

32. Vellakkal S, Gupta A, Khan Z, Stuckler D, Reeves A, Ebrahim S, et al. Has India’s national rural health mission reduced inequities in maternal health services? A pre-post repeated cross-sectional study. Health Policy Plan. 2017;32: 79–90. doi:10.1093/heapol/czw100

33. Kim R, Mejí a-Guevara I, Corsi DJ, Aguayo VM, Subramanian S V. Relative importance of 13 correlates of child stunting in South Asia: Insights from nationally representative data from Afghanistan, Bangladesh, India, Nepal, and Pakistan. Soc Sci Med. 2017. doi:10.1016/j.socscimed.2017.06.017

34. Ministry of Health and Family Welfare, Mission NH. Mothers’ Absolute Affection: Programme for Promotion of Breastfeeding. Operational Guidelines 2016. New Delhi, India; 2016. Available: http://nrhm.gov.in/MAA/Operational_Guidelines.pdf

35. Ministry of Women and Child Development. Administrative guidelines for implementation of National Nutrition Mission. New Delhi, India; 2018.

36. Ministry of Health and Family Welfare. The National Health Mission | Framework for Implementation. New Delhi, India; 2012. Available: http://nrhm.gov.in/images/pdf/NHM/NHM_Framework_for_Implementation08-01-2014_.pdf

